# Urine DNA Methylation as a Potential Diagnostic and Surveillance Tool in Non-Muscle Invasive Bladder Cancer in Thailand: A Pilot Study

**DOI:** 10.64898/2026.09.18.26363402

**Authors:** Thongtra Watcharawitayakul, Julin Ophanurak, Dutsadee Sowanthip, Poorichaya Somparn, Trairak Pisitkun, Panot Sainamthip

## Abstract

Non-muscle invasive bladder cancer (NMIBC) exhibits high recurrence, necessitating frequent invasive surveillance. Urine DNA methylation profiling represents a promising non-invasive biomarker approach for early detection and monitoring. This pilot study aimed to investigate urine DNA methylation patterns in NMIBC among Thai patients. Five patients with high-grade NMIBC and three healthy volunteers were enrolled, and midstream urine and blood samples were collected prior to transurethral resection of bladder tumor (TUR-BT). Urine cell-free DNA (cfDNA) was extracted, mechanically sheared from an atypical peak of approximately 400 bp to an optimal target length of 150–200 bp, and analyzed using cell-free methylated DNA immunoprecipitation followed by high-throughput sequencing (cfMeDIP-seq). Differentially methylated regions (DMRs) were identified between NMIBC and controls, and principal component analysis (PCA) was performed. A total of 82 DMRs were identified (adjusted *P* < 0.001), including 76 hypermethylated and 6 hypomethylated regions, with *IRX4, TBX5*, and *MIR124-3* representing the top hypermethylated genes. To account for cohort heterogeneity, a refined subset analysis excluding two outlier samples (UC2 and UC3) was performed, revealing 476 significant DMRs (449 hypermethylated). PCA demonstrated clear, robust segregation between NMIBC and controls, highlighting *MIR124-3, IRX4*, and *LHX1-DT* as the most significantly hypermethylated targets. Our findings demonstrate that urine cfDNA methylation profiling can robustly distinguish NMIBC from healthy controls, highlighting *MIR124-3, IRX4*, and *LHX1-DT* as highly promising, demographic-specific biomarkers. This non-invasive approach successfully bypasses the degradative limits of traditional bisulfite conversion and establishes a technically validated foundation for clinical surveillance and diagnostics

## Introduction

Bladder cancer represents a profound epidemiological challenge, currently ranking as the ninth most commonly diagnosed malignancy worldwide with over 614,000 new cases and 220,000 deaths reported globally in 2022. [1] While incidence rates have historically been highest in Western countries, the disease burden in Southeast Asia is rapidly escalating. In Thailand specifically, bladder cancer is a growing public health concern, currently ranking as the eighth most common cancer across all demographics and exhibiting a concerningly high mortality-to-incidence ratio. Urothelial carcinoma (UC) constitutes the vast majority of these cases, with approximately 75% to 80% initially presenting as non-muscle invasive bladder cancer (NMIBC).[2, 3]

Although NMIBC generally carries a favorable overall survival rate, it is defined by a uniquely high propensity for local recurrence—reaching up to 70% within five years—and a constant risk of progression to muscle-invasive disease.[2] This chronic relapsing nature necessitates lifelong, strict surveillance regimens mandated by international guidelines, placing an immense financial and psychological burden on patients and healthcare systems. The current standard of care relies heavily on repeated white-light cystoscopy combined with voided urine cytology.[3] However, cystoscopy is an invasive and uncomfortable procedure with inherent risks. Furthermore, urine cytology, despite its high specificity for high-grade disease, suffers from woefully inadequate sensitivity for low-grade tumors, often hovering between 30% and 40%.[4] Consequently, there is an urgent, unmet clinical need for highly sensitive, non-invasive diagnostic alternatives.[5]

Liquid biopsy utilizing urine cell-free DNA (cfDNA) has inaugurated a new frontier in urologic oncology.[5, 6] Because the urothelium is in continuous physical contact with stored urine, tumor-derived cfDNA is shed directly into the urinary tract, making urine an exceptionally enriched and anatomically precise biofluid. Among the molecular analytes detectable in urine, DNA methylation has emerged as a superior biological target.[7, 8] The epigenetic silencing of tumor suppressor genes through the hypermethylation of promoter CpG islands is a universal and early molecular hallmark of urothelial tumorigenesis, frequently preceding observable morphological changes. Diagnostic approaches targeting DNA methylation offer substantial advantages over traditional somatic mutation profiling, providing a highly robust, naturally amplified signal that is chemically stable within the harsh urinary environment.

Historically, the clinical translation of DNA methylation biomarkers was severely hampered by the destructive nature of bisulfite conversion, which degrades the already minimal quantities of cfDNA found in urine.[5] Recently, cell-free methylated DNA immunoprecipitation and high-throughput sequencing (cfMeDIP-seq) has revolutionized liquid biopsy protocols. By utilizing affinity-based enrichment without chemical degradation, cfMeDIP-seq requires ultra-low cfDNA input while preserving the natural fragment size distribution, enabling highly sensitive, genome-wide methylation profiling across various genitourinary malignancies.[9]

Despite the rapid commercial development of multi-target methylation panels for NMIBC surveillance, the vast majority have been developed and validated almost exclusively in Western cohorts.[10] Because cancer epigenetics are profoundly influenced by regional environmental exposures and distinct population genetics, there is a critical necessity to define demographic-specific epigenetic signatures. Furthermore, assessing specific epigenetic markers implicated in cellular proliferation and tumor suppression—such as *MIR124-3, IRX4*, and *LHX1-DT*—may yield highly specific diagnostic tools that reflect localized disease pathways.[11, 12]

Therefore, this pilot study aims to profile the urine cfDNA methylome of NMIBC patients in Thailand using cfMeDIP-seq. By comparing these regional methylation patterns with healthy controls, we seek to identify novel, highly specific epigenetic biomarkers that can optimize non-invasive early detection and surveillance strategies tailored to Southeast Asian populations.

## Materials (Patients) and methods

### Patient selection

Five patients with newly diagnosed bladder tumors undergoing transurethral resection of bladder tumor (TUR-BT) at King Chulalongkorn Memorial Hospital between first of July 2021 and first of December 2022 were enrolled. Inclusion criteria comprised suspected urothelial carcinoma (UC) and a non-muscle invasive bladder cancer (NMIBC) stage. Exclusion criteria included other histologies, muscle-invasive bladder cancer (MIBC), prior chemotherapy or radiation therapy, and insufficient tissue. This study was conducted as a non-randomized, observational case-control pilot study of diagnostic accuracy. All bladder cancer participants were prospectively recruited prior to undergoing routine, standard-of-care transurethral resection of bladder tumor (TUR-BT), and healthy volunteers were enrolled as non-cancer controls Three healthy volunteers were included as controls. Disease stage was classified according to the AJCC criteria. To comply with the Sex and Gender Equity in Research (SAGER) guidelines, biological sex was categorized based on sex assigned at birth.

### Specimen collection

Midstream urine (100 mL) was collected from patients prior to TUR-BT, supplemented with Cell-Free DNA Urine Preserve, and stored at 4°C. Blood was collected for peripheral blood mononuclear cell (PBMC) isolation. Tumor tissues were immediately preserved in liquid nitrogen.

### cfDNA extraction and cfMeDIP-seq

Urine cfDNA was extracted using the QIAamp Circulating Nucleic Acid Kit (QIAGEN, [Hilden, Germany]) and quantified with a Qubit fluorometer (Invitrogen, [Carlsbad, CA, USA]). Because initial evaluations revealed cfDNA fragment lengths of approximately 400 bp, the DNA was mechanically sheared to an optimal target length of 150–200 bp prior to library preparation. cfMeDIP-seq libraries were prepared using the KAPA HyperPrep Kit (KAPA Biosystems, [Wilmington, MA, USA]) and NEBNext adaptors. Immunoprecipitation was performed with the MagMeDIP kit (Diagenode, [Seraing, Belgium]) and quality-controlled using spiked-in methylated and unmethylated *Arabidopsis thaliana* filler DNA. Libraries were subsequently sequenced on an Illumina HiSeq 4000 platform (150-bp paired-end; Illumina, [San Diego, CA, USA]).

### Data analysis

Sequence reads were trimmed using Trim Galore and aligned to the human reference genome (hg38) using Bowtie 2. Duplicate reads were sorted and removed using SAMtools. Differentially methylated regions (DMRs) were identified using edgeR (bin size: 300) with an adjusted *P*-value threshold, and genomic annotation was performed using HOMER. Finally, principal component analysis (PCA) was conducted to visualize methylation patterns and sample clustering

### Ethics statement

The study was conducted in accordance with the Declaration of Helsinki, and the study protocol was approved by the Institutional Review Board of the Faculty of Medicine, Chulalongkorn University (Approval No. COA No.979/2021 and IRB No.RA64/053). All participants gave informed written consent for study participation prior to inclusion. This study was registered with Thai Clinical Trials Registry number TCTR20210813003.

## Results

### Patient characteristics and urine cfDNA profiles

A total of eight participants were included in this sequencing cohort: five patients with newly diagnosed NMIBC and three healthy controls. The NMIBC cohort consisted of four males and one female, with a median age of 76 years (range 57–87 years). Based on pathological evaluation, the tumor stages were Ta (n=3) and T1 (n=2), and notably, all five tumors were classified as high-grade urothelial carcinoma.(Table1) Quantitative analysis of urine cell-free DNA (cfDNA) revealed that healthy controls secreted significantly lower concentrations of cfDNA compared to NMIBC patients. Interestingly, initial fragment size analysis demonstrated that the extracted urine cfDNA possessed an unusually large peak size of approximately 400 bp, which is notably longer than standard apoptotic cfDNA fragments. Consequently, the cfDNA was mechanically sheared to an optimal target length of 150–200 bp prior to downstream library preparation to ensure sequencing efficiency

**Table 1.**
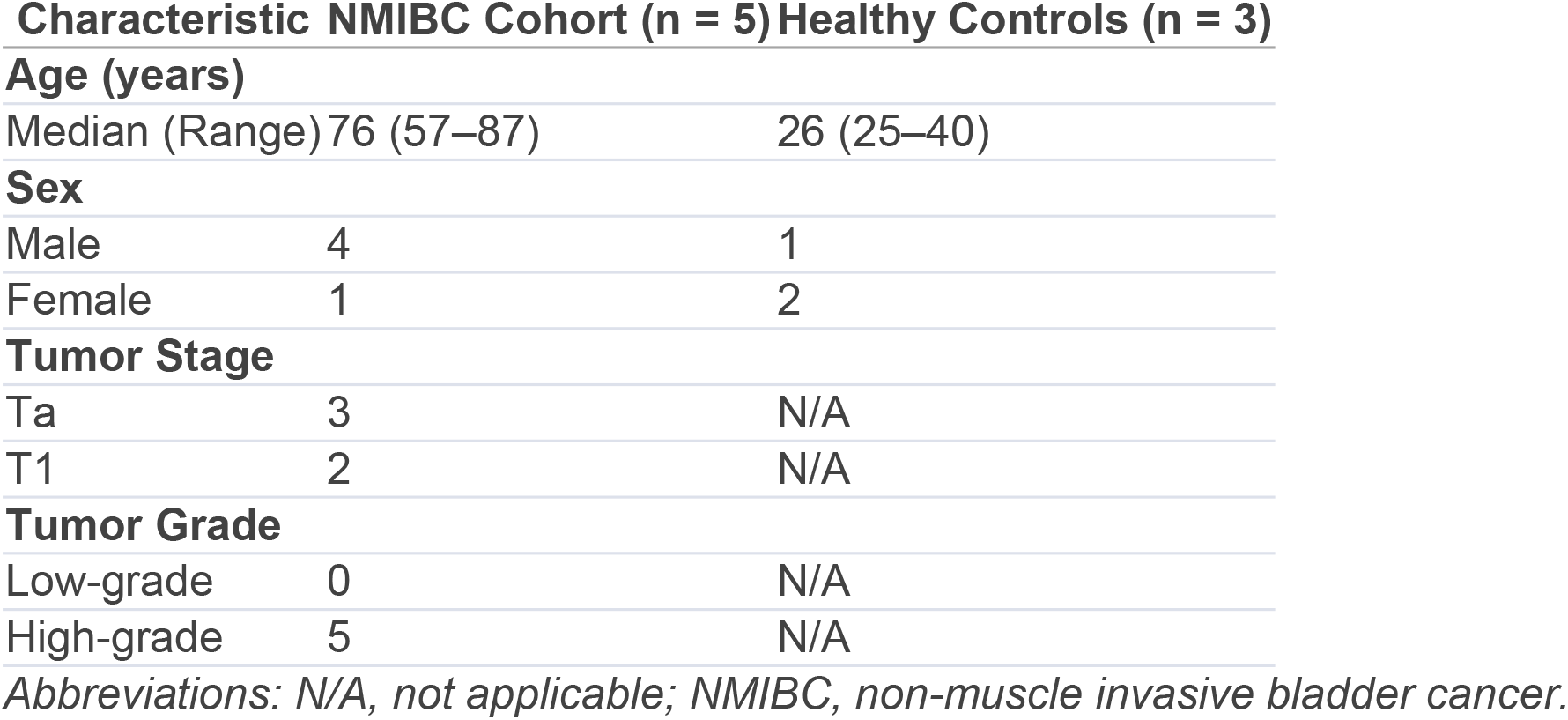
Baseline patient and sample characteristics.

### Quality control of cfMeDIP-seq data

Following sequencing, an average of 45 million paired-end reads per sample were obtained. Alignment to the hg38 reference genome yielded a high unique mapping rate, and the duplication rate remained strictly <5% across all samples. Quality control utilizing spiked-in methylated and unmethylated *Arabidopsis thaliana* DNA demonstrated >99% recovery of methylated DNA and <1% recovery of unmethylated DNA, confirming the high specificity and success of the immunoprecipitation process

### Quality control of cfMeDIP-seq data

Following sequencing, an average of 45 million paired-end reads per sample were obtained. Alignment to the hg38 reference genome yielded a high unique mapping rate, and the duplication rate remained strictly <5% across all samples. Quality control utilizing spiked-in methylated and unmethylated *Arabidopsis thaliana* DNA demonstrated >99% recovery of methylated DNA and <1% recovery of unmethylated DNA, confirming the high specificity and success of the immunoprecipitation process

### Genome-wide methylation profiling of the complete NMIBC cohort

Comparing the complete NMIBC cohort (n=5) to the healthy controls (n=3), we identified 82 significant differentially methylated regions (DMRs) (adjusted *P* < 0.001), comprising 76 hypermethylated and 6 hypomethylated regions. Genomic annotation of these significant DMRs revealed that they were predominantly localized to intergenic regions (41%), introns (29%), exons (15%), promoter-TSS regions (11%), and 5’ UTRs (4%). Furthermore, principal component analysis (PCA) demonstrated a clear segregation of global methylation profiles between the complete NMIBC patient cohort and the healthy controls **(Fig 1A and 1B)**.

**Fig 1.**
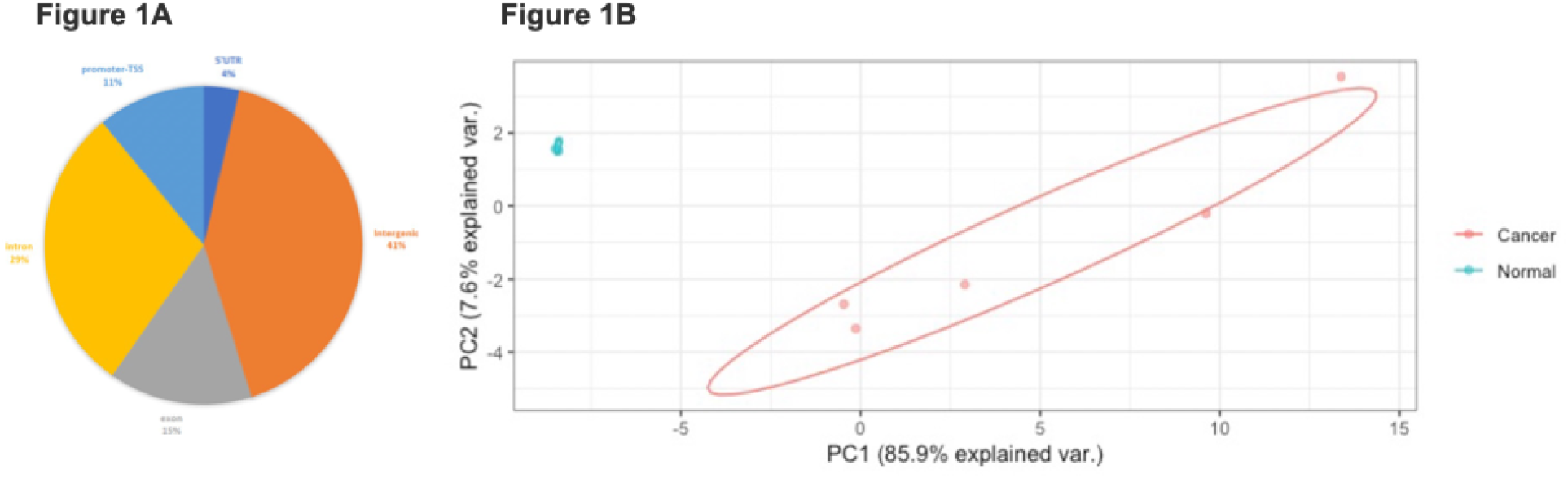
Differentially methylated regions (DMRs) in the complete non-muscle invasive bladder cancer (NMIBC) cohort compared to healthy controls. **(A)** Genomic distribution of the 82 significant DMRs, highlighting predominant localization in intergenic and intronic regions. **(B)** Principal component analysis (PCA) demonstrating clear segregation of methylation profiles between NMIBC patients and healthy controls.

### Refined subset analysis and identification of biomarker candidates

Due to the observed epigenetic heterogeneity in two tumor samples (UC2 and UC3), a refined subset analysis excluding these outliers was performed to capture the most consistent tumor-specific signatures. In this refined subset, 476 significant DMRs were identified (449 hypermethylated, 27 hypomethylated; adjusted *P* < 0.001). The genomic distribution in this subset slightly shifted to: introns (29%), intergenic regions (24%), exons (20%), promoter-TSS regions (17%), 5’ UTRs (5%), non-coding regions (3%), 3’ UTRs (1%), and TTS (1%). Principal component analysis (PCA) and hierarchical clustering of this refined subset confirmed a robust separation between the NMIBC samples and controls **(Fig 2A, 2B, and 2C)**. Gene-level analysis highlighted several novel targets exhibiting significant hypermethylation in the Thai NMIBC cohort. The highest magnitudes of hypermethylation were consistently observed in *MIR124-3, IRX4, TBX5, LHX1-DT, TAFA2*, and *HSF4*. These densely hypermethylated loci present highly promising, non-invasive biomarker candidates for the early detection and surveillance of high-grade NMIBC.

**Fig 2.**
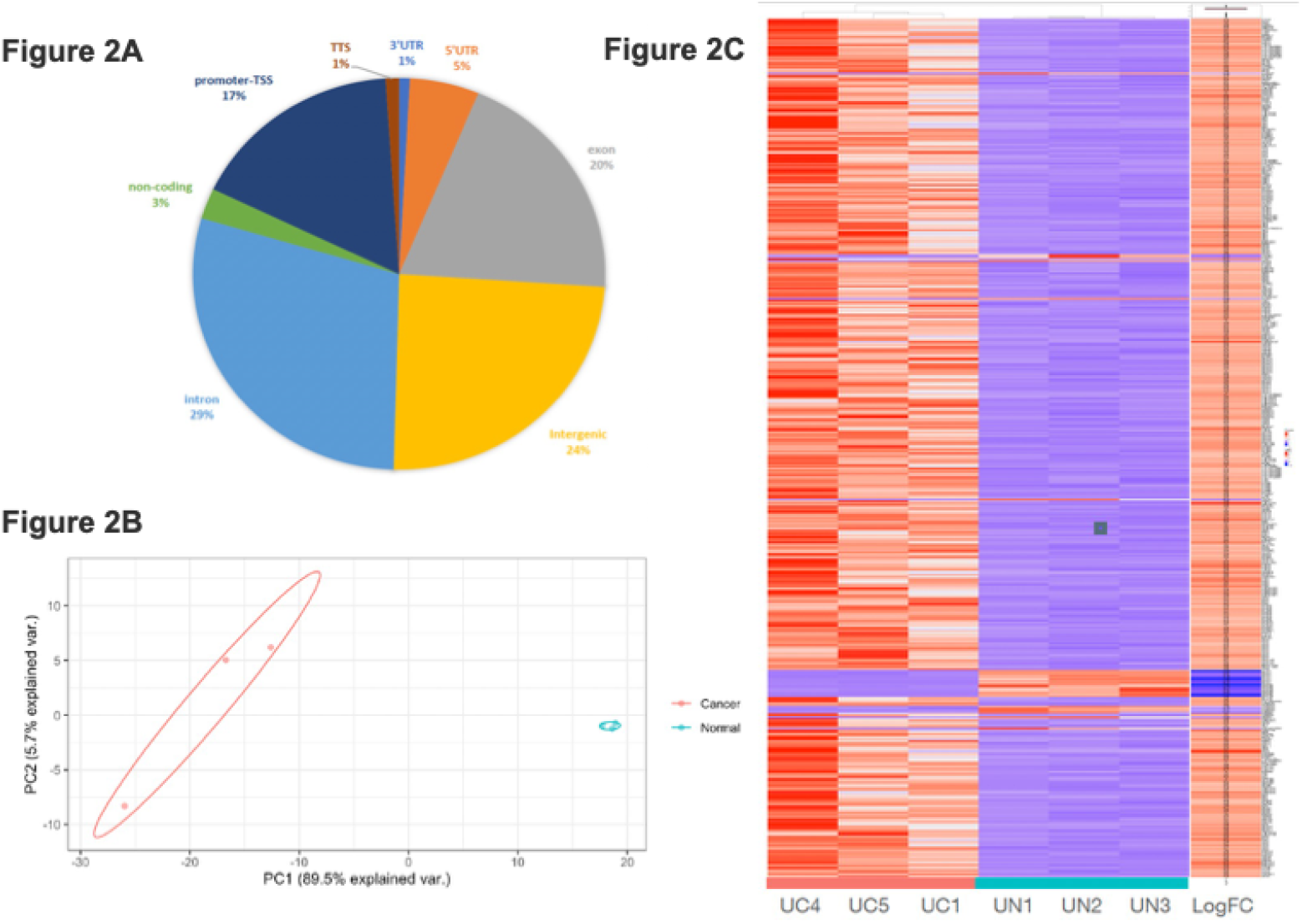
Refined subset analysis of genome-wide methylation patterns. **(A)** Genomic distribution of the 476 significant DMRs identified following the exclusion of highly heterogeneous outlier samples. **(B)** Principal component analysis (PCA) and **(C)** hierarchical clustering heatmaps demonstrating robust separation between NMIBC and controls, highlighting *MIR124-3, IRX4*, and *LHX1-DT* as the most significantly hypermethylated targets. *Abbreviations: DMR, differentially methylated region; NMIBC, non-muscle invasive bladder cancer; PCA, principal component analysis*.

## Discussion

Epigenetic profiling, specifically aberrant DNA methylation at CpG islands, has been extensively established as a universal and early molecular hallmark of urothelial tumorigenesis, frequently preceding observable morphological changes. While multiple urinary biomarker panels are commercially available—such as the 15-marker Bladder EpiCheck test, which has demonstrated a negative predictive value of up to 98% for high-grade disease[13, 14]—their widespread clinical adoption has been limited. The vast majority of these multi-target panels were developed and validated almost exclusively in Western cohorts. Because cancer epigenetics are profoundly influenced by regional environmental exposures and distinct population genetics [15], identifying demographic-specific epigenetic signatures is a critical necessity.

Our pilot data successfully identified a highly distinct set of hypermethylated loci—*MIR124-3, IRX4*, and *LHX1-DT*—in the urine cfDNA of Thai NMIBC patients. The identification of these specific targets aligns deeply with the fundamental oncogenic pathways driving urothelial carcinoma. For instance, *MIR124-3* functions as a potent tumor-suppressive microRNA.[16, 17] Its epigenetic silencing abolishes the post-transcriptional suppression of ROCK1 and STAT3[18], thereby hyper-driving actin-myosin contractility, cell migration, and uncontrolled cell proliferation. Detecting *MIR124-3* hypermethylation in urine cfDNA thus effectively captures the molecular preconditions for tumor invasion. Similarly, *IRX4* acts as a vital epithelial tumor suppressor gene that regulates the PI3K/AKT and ERK signaling pathways through downstream targets like *CRYAB*. Promoter hypermethylation of *IRX4* systematically removes this vital transcriptional checkpoint, accelerating cancer cell growth.[11, 19] Furthermore, *LHX1-DT*, a long non-coding RNA, has been strongly implicated in actively promoting cancer cell proliferation and evasion of apoptosis in renal cancer.[20] The dense hypermethylation of these three loci in our NMIBC cohort strongly supports their role in localized disease pathogenesis and surveillance.

Compared with existing urine methylation panels, our highly specific localized epigenetic signature potentially reflects population-specific patterns unique to Southeast Asian demographics. Historically, the clinical translation of evaluating DNA methylation from urine was severely hampered by the destructive nature of bisulfite conversion, which degrades up to 90% of the already minimal quantities of urinary cfDNA.[21] By utilizing cfMeDIP-seq, our study decisively bypassed this limitation. cfMeDIP-seq is highly sensitive, requires ultra-low cfDNA input (often <10 ng), relies on non-destructive affinity-based enrichment, and preserves the natural fragment size distribution, thereby enabling comprehensive and unbiased genome-wide methylation profiling.[6]

Several important limitations of this pilot study must be acknowledged. First, the sample size is inherently small (n = 5 NMIBC cases, n = 3 healthy controls) and derived from a single institution, which limits the statistical power and broader generalizability of the findings. Second, our NMIBC cohort exclusively consisted of patients with high-grade urothelial carcinoma. Because low-grade tumors typically shed less tumor DNA and often harbor fewer epigenetic aberrations compared to high-grade disease, the sensitivity and diagnostic performance of our *MIR124-3, IRX4*, and *LHX1-DT* panel for detecting low-grade NMIBC remains completely unassessed. Third, there are significant technical and pre-analytical challenges regarding urine liquid biopsies. As observed in our methodology, the extracted urinary cfDNA exhibited unexpected fragment length variations—peaking at approximately 400 bp—which necessitated additional mechanical shearing prior to library preparation to ensure sequencing efficiency. Finally, while cfMeDIP-seq provides exceptional, unbiased genome-wide profiling without the destructive effects of bisulfite conversion, the technique currently requires extensive bioinformatic infrastructure, has a lengthy turnaround time, and is not yet cost-effective or practical for routine, high-throughput clinical surveillance in a standard hospital setting.

Despite these limitations, the clear principal component segregation of NMIBC samples from controls underscores the biological validity of this epigenetic approach. Larger, multi-center validation studies with longitudinal follow-up—specifically incorporating low-grade NMIBC cohorts—are warranted. Furthermore, translating these genome-wide sequencing discoveries into rapid, targeted, and highly affordable clinical assays (such as droplet digital PCR or targeted multiplex panels) will be essential to establish the true clinical utility, reproducibility, and recurrence-predictive value of the *MIR124-3, IRX4*, and *LHX1-DT* panel in Asian populations.

## Conclusion

Urine cfDNA methylation profiling via cfMeDIP-seq can robustly distinguish NMIBC patients from healthy controls, effectively circumventing the historical limitations of low-yield urinary DNA analysis. *MIR124-3, IRX4*, and *LHX1-DT* emerge as highly promising, population-specific epigenetic biomarkers for Thai cohorts. Non-invasive urine liquid biopsy facilitates a patient-friendly surveillance paradigm with the potential to safely de-intensify routine invasive cystoscopy intervals and proactively forecast disease recurrence. Further large-scale, longitudinal studies are required to fully validate these specific markers for clinical integration in Southeast Asian populations.

## Data Availability

The raw sequencing datasets (cfMeDIP-seq) generated and analyzed during the current study have been deposited in the NCBI Sequence Read Archive (SRA) and will be provided upon manuscript acceptance. All other minimal clinical and baseline demographic data necessary to replicate the findings are included within the manuscript and its Supporting Information files.

## Abbreviations

NMIBC: Non-muscle invasive bladder cancer
MIBC: Muscle-invasive bladder cancer
TURBT: Transurethral resection of bladder tumor
cfDNA: Cell-free DNA
cfMeDIP-seq: Cell-free methylated DNA immunoprecipitation and high-throughput sequencing
DMRs: Differentially methylated regions
PCA: Principal component analysis.

## Acknowledgements

We would like to thank the staff at King Chulalongkorn Memorial Hospital and the Systems Biology Center for their technical support.

## Authors’ contributions

TW, PS, JO, DS, PuS, and TP contributed to the study concept and design. Data acquisition was performed by TW, PS, JO, DS, PuS, and TP. Data analysis was performed by TW and PS. The first draft of the manuscript was written by TW and PS. All authors critically revised the manuscript. All authors read and approved the final version of the manuscript.

## Consent for publication

Not applicable.

## References

1. Bray F, Laversanne M, Sung H, Ferlay J, Siegel RL, Soerjomataram I, Jemal A: Global cancer statistics 2022: GLOBOCAN estimates of incidence and mortality worldwide for 36 cancers in 185 countries. CA Cancer J Clin 2024, 74(3):229–263.

2. Chang SS, Boorjian SA, Chou R, Clark PE, Daneshmand S, Konety BR, Pruthi R, Quale DZ, Ritch CR, Seigne JD et al: Diagnosis and Treatment of Non-Muscle Invasive Bladder Cancer: AUA/SUO Guideline. J Urol 2016, 196(4):1021–1029.

3. Gontero P, Birtle A, Capoun O, Comperat E, Dominguez-Escrig JL, Liedberg F, Mariappan P, Masson-Lecomte A, Mostafid HA, Pradere B et al: European Association of Urology Guidelines on Non-muscle-invasive Bladder Cancer (TaT1 and Carcinoma In Situ)-A Summary of the 2024 Guidelines Update. Eur Urol 2024, 86(6):531–549.

4. Yafi FA, Brimo F, Steinberg J, Aprikian AG, Tanguay S, Kassouf W: Prospective analysis of sensitivity and specificity of urinary cytology and other urinary biomarkers for bladder cancer. Urol Oncol 2015, 33(2):66 e25–31.

5. Sun X, Wang D, Zhang S, Wang J, Ning H, Wu H, Wu F, Tang D, Lyu J: Unleashing the potential of urine DNA methylation detection: Advancements in biomarkers, clinical applications, and emerging technologies. Curr Urol 2025, 19(5):295–302.

6. Chaudhuri AA, Pellini B, Pejovic N, Chauhan PS, Harris PK, Szymanski JJ, Smith ZL, Arora VK: Emerging Roles of Urine-Based Tumor DNA Analysis in Bladder Cancer Management. JCO Precis Oncol 2020, 4.

7. Kandimalla R, van Tilborg AA, Zwarthoff EC: DNA methylation-based biomarkers in bladder cancer. Nat Rev Urol 2013, 10(6):327–335.

8. Harsanyi S, Novakova ZV, Bevizova K, Danisovic L, Ziaran S: Biomarkers of Bladder Cancer: Cell-Free DNA, Epigenetic Modifications and Non-Coding RNAs. Int J Mol Sci 2022, 23(21).

9. Nuzzo PV, Berchuck JE, Korthauer K, Spisak S, Nassar AH, Abou Alaiwi S, Chakravarthy A, Shen SY, Bakouny Z, Boccardo F et al: Detection of renal cell carcinoma using plasma and urine cell-free DNA methylomes. Nature Medicine 2020, 26(7):1041–1043.

10. Witjes JA, Morote J, Cornel EB, Gakis G, van Valenberg FJP, Lozano F, Sternberg IA, Willemsen E, Hegemann ML, Paitan Y et al: Performance of the Bladder EpiCheck Methylation Test for Patients Under Surveillance for Non-muscle-invasive Bladder Cancer: Results of a Multicenter, Prospective, Blinded Clinical Trial. Eur Urol Oncol 2018, 1(4):307–313.

11. Chakma K, Gu Z, Abudurexiti Y, Hata T, Motoi F, Unno M, Horii A, Fukushige S: Epigenetic inactivation of IRX4 is responsible for acceleration of cell growth in human pancreatic cancer. Cancer Sci 2020, 111(12):4594–4604.

12. Yang X, Li J, Wang Y, Li P, Zhao Y, Duan W, Ariston Gabriel AN, Chen Y, Mao H, Wang Y et al: Individualized Prediction of Survival by a 10-Long Non-coding RNA-Based Prognostic Model for Patients With Breast Cancer. Front Oncol 2020, 10:515421.

13. Cano Velasco J, Artero Fullana S, Polanco Pujol L, Lafuente Puentedura A, Subiela JD, Aragon Chamizo J, Moralejo Garate M, Hernandez Fernandez C: Use of Bladder Epicheck(R) in the follow-up of non-muscle-invasive Bladder cancer: A systematic literature review. Actas Urol Esp (Engl Ed) 2024, 48(8):555–564.

14. Palermo M, D’Elia C, Trenti E, Comploj E, Mian C, Schwienbacher C, Heidegger I, Clauser S, Pycha A, Vjaters E: Prospective evaluation of the RT-PCR based urinary marker Bladder Epicheck(R) as a diagnostic tool in upper urinary tract tumor. Minerva Urol Nephrol 2024, 76(2):195–202.

15. Guerrero-Bosagna C, Skinner MK: Environmentally induced epigenetic transgenerational inheritance of phenotype and disease. Mol Cell Endocrinol 2012, 354(1–2):3–8.

16. Patil N, Abba ML, Zhou C, Chang S, Gaiser T, Leupold JH, Allgayer H: Changes in Methylation across Structural and MicroRNA Genes Relevant for Progression and Metastasis in Colorectal Cancer. Cancers (Basel) 2021, 13(23).

17. Loginov VI, Burdennyy AM, Filippova EA, Pronina IV, Lukina SS, Kazubskaya TP, Karpukhin AV, Khodyrev DS, Braga EA: Aberrant Methylation of 21 MicroRNA Genes in Breast Cancer: Sets of Genes Associated with Progression and a System of Markers for Predicting Metastasis. Bull Exp Biol Med 2021, 172(1):67–71.

18. Kayisaier K, Abulajiang T, Tang L, Hasiyeti W, Gulibositan M: [Effect of miR-340-5p on proliferation of laryngeal cancer Hep2 cells and its intrinsic molecular mechanism]. Lin Chuang Er Bi Yan Hou Tou Jing Wai Ke Za Zhi 2020, 34(2):140–145.

19. Jia M, Kong D, Fan J, Heng B, Zhao J, Li B, Liu S, Kong P, Liu Y: Anti-colorectal cancer effects of IRX4 and sensitivity studies to oxaliplatin. Front Immunol 2025, 16:1581244.

20. Zhu C, Li R, You X, Xu J, Wang J, Dong D, Chen X, Wang K: m6A reader IGF2BP2-stabilized lncRNA LHX1-DT inhibits renal cell carcinoma (RCC) cell proliferation and invasion by sponging miR-590-5p. NPJ Precis Oncol 2025, 9(1):193.

21. Larsen LK, Lind GE, Guldberg P, Dahl C: DNA-Methylation-Based Detection of Urological Cancer in Urine: Overview of Biomarkers and Considerations on Biomarker Design, Source of DNA, and Detection Technologies. Int J Mol Sci 2019, 20(11).

